# RURAL, LOW-INCOME, HIGH-ALTITUDE POPULATION: BURDEN, DETERMINANTS OF POSTDATE PREGNANCIES AND NEONATAL PHENOTYPE

**DOI:** 10.1101/2023.06.14.23291386

**Authors:** Adenike Oluwakemi Ogah, James Aaron Ogbole

**Author notes:** **Corresponding author:** Dr Adenike Oluwakemi Ogah, PhD, PhD.

## Abstract

**Background:** Conditions, which reduce oxygen delivery to the fetus, are associated with impaired intrauterine growth. Postdate babies are thought to be rare in today’s neonatal practice. The combined effects of prolonged gestation, high altitude and rural-low-income residence on fetal growth and hence timing of delivery, have been poorly investigated. This study examined and compared the burden, determinants and neonatal phenotypes of term and postdate pregnancies in a remote, high-altitude (1674m above sea level) community.

**Subject and methods:** This was the secondary analysis of the baseline data collected for a prospective cohort study. Healthy, 529 mother-singleton infant pairs were recruited consecutively from a rural district hospital. Newborns were classified by their gestational age and weight according to WHO. Gestational age was determined by mother’s LMP, fetal ultrasound dating and post-delivery ballard examination. Postdate was defined by gestation >40weeks. Preterm babies (26-4.9%) were excluded, remaining with 503 babies that were used in the analysis. Logistic regression models and chi test were used to compare the characteristics of term and postdate deliveries.

**Results:** Of the 503 mother-infant study participants, postdate babies were 199 (39.6%) and SGA babies were 108 (21.4%). Overall, SGA birth rate was 4 times higher than the preterm birth rate. There was greater odds of SGA birth (p<0.001; OR=11.05; 95% CI 3.74, 35.57), especially symmetrically growth retarded SGA (p=0.003; OR=6.60; 95% CI 1.67, 26.1) in postdate delivery compared to term delivery. Though birthweight was preserved, the rate of SGA delivery progressively increased from 14.3% at 38weeks to 80% at 43weeks gestation. The rate of AGA delivery peaked at 40 weeks gestation (85.6%) and thereafter declined. Maternal married status (p=0.009, OR=2.27) and high MUAC (p=0.049; OR =1.74) promoted postdate pregnancy. Mothers (80%, p=0.005) engaged in professional occupation had a tendency to prolonged pregnancy. Good environmental (water and toilet) sanitation was more likely to result in postdate delivery, p<0.05.

**Conclusion and Recommendations:** The rates of postdate and SGA births were unexpectedly high for the level of altitude. Unlike in low land studies, where prolonged pregnancies were associated with macrosomia, postdate deliveries in this high-altitude population, were more commonly associated with SGA birth and their complications. The strong association of symmetric SGA (rather than asymmetric SGA) with postdate pregnancies suggests longstanding intrauterine insult from probably combined effects of chronic hypoxia from high altitude residence and maternal factors. The authors of this study strongly recommend that high-altitude pregnancies should not be allowed to progress beyond 40weeks gestation and more resources should be committed to identifying high risk pregnancies with tendencies for prolonged pregnancies and improve maternal nutrition and oxygen status during pregnancy. Newborns should be classified at birth, their problems anticipated and managed appropraitely for an improved neonatal care, survival and wellbeing.

## Background

On the average, gestation lasts 40 weeks (280 days) from the first day of the last menstrual period to the estimated date of delivery in singleton pregnancies.^1^ Previously, the period from 37 weeks to 42 weeks was considered “term,” with the expectation that neonatal outcomes from deliveries during this interval, would be uniform and good. However, research has increasingly revealed that neonatal outcomes, particularly respiratory morbidity, is unpredictable, depending on the timing of delivery, even within this five-week gestational age range.^2^ Newer definitions of gestation include: term (37 0/7-40 6/7 weeks of gestation), late term (41 0/7-41 6/7 weeks of gestation) and postterm (≥ 42weeks gestation). Though, the cut-off points vary with different definitions, that of postterm gestation has remained consistent.^3^ The frequency of postterm pregnancy was estimated to be 3-12%. The actual rate might be lower, because the most frequent cause of postterm pregnancy is inaccurate dating.^3^

The risk of adverse neonatal outcomes is lowest in uncomplicated pregnancies delivered between 39 0/7 and 40 6/7 weeks of gestation.^4^ The adverse risks to the mother and fetus increase after 41 weeks, mainly due to increasing fetal weight; decline in placental function; fetal hypoxia; oligohydramnios, which increase chances of cord compression and meconium aspiration. Perinatal mortality after 42 weeks is twice as compared to the perinatal mortality at 40 weeks and by 44 weeks the rate is increased up to threefold.^4,5,6^

There is a debate on the optimum time to induce labor in low-risk mothers, whether at 41 weeks + 0 days or to allow the pregnancy to continue until 42 weeks + 0 days. For mothers living in rural, low-income, high-altitude locations, poor fetal outcome might actually set in much earlier than expected and therefore induction of labour might need to be considered much earlier. Unfortunately, there is paucity of literature in this aspect.

According to Ranjbar et al. (2023)’s report (involving 8,888 singleton deliveries, preterm and postterm babies were excluded), late-term pregnancies had a higher risk of macrosomia, meconium amniotic fluid and fetal distress. When compared to term pregnancy, the risk of low birth weight (LBW) was lower in late-term pregnancies. Ranjbar’s study was carried out in Bandar Abbas city (urban area, with elevation of 9m above sea level), Iran between January 2020 and 2022.^7^ Kandalgaonkar et al. in a prospective cross-sectional study of 5110 participants in India, prevalence of postdate pregnancies was 5.1% and there was increased use of c-section delivery in postdated pregnancies compared to term pregnancies.

High altitude pregnancies have been more commonly investigated for preterm births rather than postdate delivery in several literature.^5^Chronic hypoxia, associated with living at high altitude, reduces uterine artery blood flow, slows fetal growth, therefore resulting in fetal growth retardation.^8^ Fetal hypoxia and complications are further compounded in prolonged pregnancy resident at high altitude. Grant et al. (2021) observed, in a systematic review involving 59 studies and 1, 604, 770 pregnancies, that high altitude pregnancies are at increased risk for LBW, SGA, and spontaneous preterm birth.^9^ Maternal genetic factors is known to account for about 30% of these postdate pregnancies.^10^ Other risk factors for postterm pregnancy include primiparity, previous postterm pregnancy,^11^ male fetus,^12^ obesity.^13^

Little is known about the link between postdate pregnancies (beyond 40 weeks gestation) and poor neonatal outcomes in rural, low-income, high-altitude settings. In this study, we aim to determine the prevalence of and factors associated with postdate deliveries and their neonatal outcomes in this peculiar population. This study will further inform and guide decisions made by health care policy makers, clinicians and other stakeholders on the most appropriate time to induce labor for a better neonatal outcome, especially in rural high-altitude settings.

## Concept of the study

Figure 1, shows the maternal, neonatal and environmental predictors of postdate and term deliveries (primary outcome), that were investigated in this study. The secondary outcomes were SGA (symmetric/asymmetric SGA), breastfeeding initiation interval and prelacteal feeding.

**Fig 1:**
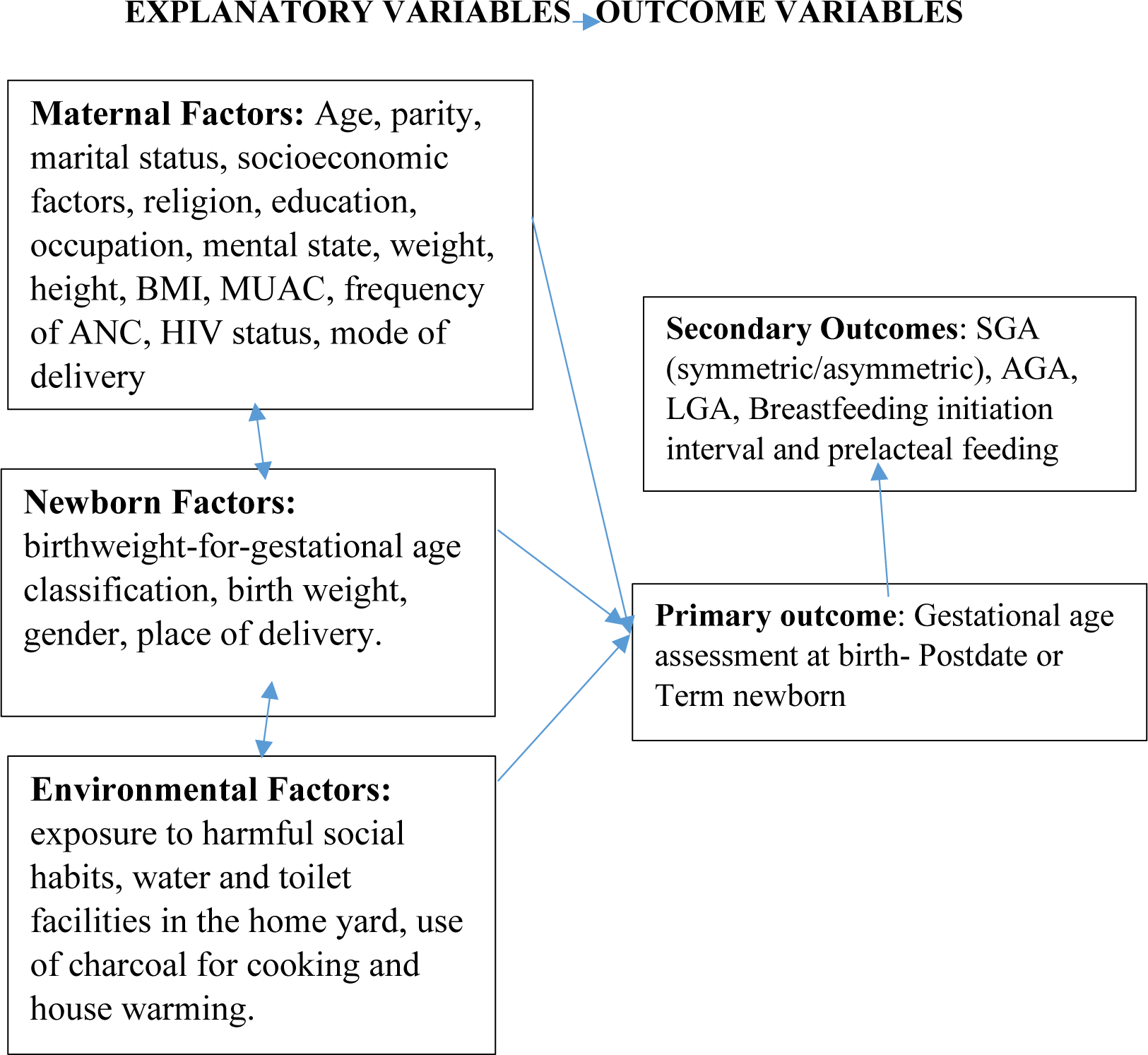
Concept of the study.

## Materials and methods

The methods employed in carrying out this study is discussed in this section.

### Study setting

Rwanda is a low-income, agricultural and landlocked country with approximately 11 million people living in five provinces, covering an area of 26,338 km^2^. It is called the home of a ‘1000 hills’.^14,15,16^ The limited area of flat land available in most part of Rwanda is a hindrance to farming, animal rearing and construction of standard residential houses, among others. For example, the recommended minimum of 50 feet distance between source of household water and sewage tank in residential yards are often compromised during construction, leading to contamination of water source. There are 2 peak raining seasons in the country: April to June and September to November. Non-availability of regular supplies of clean and safe water, has been a longstanding problem in Rwanda, as a whole, probably because of its landlocked and hilly terrains, making construction and supply of piped water, a major challenge. The flow of water in public pipes is infrequent and the taps and pipes may be rusted and breached in some places, especially in the rural areas, further leading to contamination of household water. Many families store rain water in big tanks for use in their homes and this may become polluted (in the writer’s opinion) because of difficulties of cleaning these storage tanks and their proximity to sewage tanks situated in the home yards. A few non-profit organizations, such as USAID and Water-for-life have sunk bore holes in strategic locations in a few number of villages in the country, with the aim of alleviating this water problem.^14,15^

Rwanda has an average of 4.4 persons per household^15^ and a gross domestic product per capita of US $780.80.^15^ About half (48%) of its population is under 19 years of age and 39% live below the poverty line with a life expectancy at birth of 71.1 years for women and an adult literacy rate of 80% among 15–49 years old women. In addition, 87.3% of the population have health insurance and access to health services; spending an average of 47.4 min to reach a health centre.^15^

Gitwe village (current study site) is located on a high altitude of 1,674 meters above sea level, in the southern province, 240km from Kigali, which is the capital city of Rwanda. Gitwe general hospital began in 1995, immediately after the genocide, for the purposes of providing medical services and later training to this isolated community. The hospital currently has 100% government support, since year 2020. The maximum number of deliveries at the hospital per month was about 200. Some of the challenges in the hospital include poor specialist coverage and few trained health workers, poor supply of equipment, water, electricity, laboratory services and medicines.

Challenging cases are referred to the University of Rwanda Teaching Hospital in Butare or Kigali. Gitwe village was selected for this study, because there was no published birth data on this poorly researched, remote community. In 2019, birth, feeding and growth data on 529 healthy mother-singleton newborn pairs were compiled by the investigator, over a period of 12 months in the delivery and postnatal wards of Gitwe general hospital and at its annex, the maternal and child health clinic.

### Data source and sample

This was a secondary analysis of the baseline data collected for a prospective cohort study in 2019. Mother-healthy singleton newborn pairs were recruited consecutively, on first-come-first-serve basis. Maternal file review and newborn anthropometry [weight (kg), length (cm) and head circumference (cm) measurements, recorded to the nearest decimals] were carried out, soon after birth. Maternal sociodemographic, antenatal and nutritional data were obtained using questionnaires, which were read to the mothers and filled by the research assistants. Maternal anthropometry was measured at 6 weeks postpartum and recorded and their BMI and MUAC were computed. Three approaches were used to classify the newborn: gestational age (in completed weeks), birthweight and birthweight-gestational age percentile (WHO). Newborn gestational age was determined using the maternal last menstrual period (LMP), fetal ultrasound gestational age dating (preferably done at first trimester) and/or expanded new Ballard criteria and were classified into preterm (gestation age <37weeks), term (gestational age between 37-40 weeks) and post-date (gestational age >40weeks). Birthweight regardless of gestational age, with cut-off points of 2.5-3.9kg for normal birth weight. Birthweight-gestational age newborn classification (globally recommended) into small-for-gestational age (SGA), appropriate-for-gestational age (AGA) and large-for-gestational age (LGA) was done: according to WHO using 10^th^-90^th^ percentiles cut-off points for AGA. The SGA babies were further classified into symmetrical (PI≥2) and asymmetrical (PI<2), based on their Ponderal’s index. The Ponderal’s index was calculated thus: PI = Weight (GM)/Length (CM) 3 × 100.^18^ 26 (4.9%) preterm babies were excluded from the study analysis, remaining with 503 eligible mother-newborn study participants. It is worth noting that newborn classification is rarely done in the study site. Newborn place and mode of delivery and environmental conditions were also recorded.

To ensure the quality of data collected, 2 registered nurses were trained as research assistants at Gitwe Hospital for 2 days on the over-all procedure of mother and newborn anthropometry and data collection by the investigator. The questionnaires were pre-tested before the actual data collection period, on 10 mother-infant pair participants (2% of the total sample). The investigator closely followed the day-to-day data collection process and ensured completeness and consistency of the questionnaires administered each day, before data entry.

### Statistical analysis

Data clean up, cross-checking and coding were done before analysis. These data were entered into Microsoft Excel statistical software for storage and then exported to SPSS version-26 for further analysis. Both descriptive and analytical statistical procedures were utilized. Participants’ categorical characteristics were summarised in frequencies and percentages. The chi-squared test was used to compare the categorical differences between the postdate and term groups. Binary logistic regression models were used to assess the associations between neonatal, maternal and environmental factors with postdate/term pregnancy. Multivariate logistic regression analysis was used to investigate the relationship between SGA (symmetrical/asymmetrical), AGA, LGA births and postdate/term births. Independent variables were maternal (age, parity, marital status, religion, social habits, education, years of schooling, occupation, weight, height, BMI, MUAC, frequency of ANC, HIV status, socioeconomic status and type of home/marraige); neonatal factors (gender, intrauterine growth and birth size); and environmental factors (use of water and toilet facilities, charcoal and harmful exposures). Factors with p-values <0.1 were fed into the regression models. Odds ratio (OR), with a 95% confidence interval (CI) were computed to assess the strength of association between independent and dependent variables. For all, statistical significance was declared at p-value < 0.05. The reporting in this study were guided by the STROBE guidelines for observational studies.^17^

### Ethics

Ethical approval from the Health Sciences Research Ethics Committee of the University of the Free State in South Africa (Ethical Clearance Number: UFS-HSD2018/1493/2901) was obtained. Written permission to collect data was obtained from the Director of Gitwe Hospital and the eligible mothers gave their informed consent before enrolment. The participants were given research identity numbers and the principal investigator was responsible for the safe keeping of the completed questionnaires and collected data, to ensure anonymity and confidentiality of the participants.

## Results

The following are the results obtained from the study.

### Participants

Five hundred and ninety-seven (597) babies were delivered at Gitwe District Hospital, Rwanda, between 3^rd^ January and 9^th^ May 2019, out of which, eligible 529 mother-newborn pairs were transferred to the postnatal ward, Figure 2. For this current study, out of the 529 babies transferred to the postnatal ward, the preterm babies (26, 4.9%) were excluded during analysis. Hence, 503 healthy mother-infant pairs were the study population for this study.

**Fig 2:**
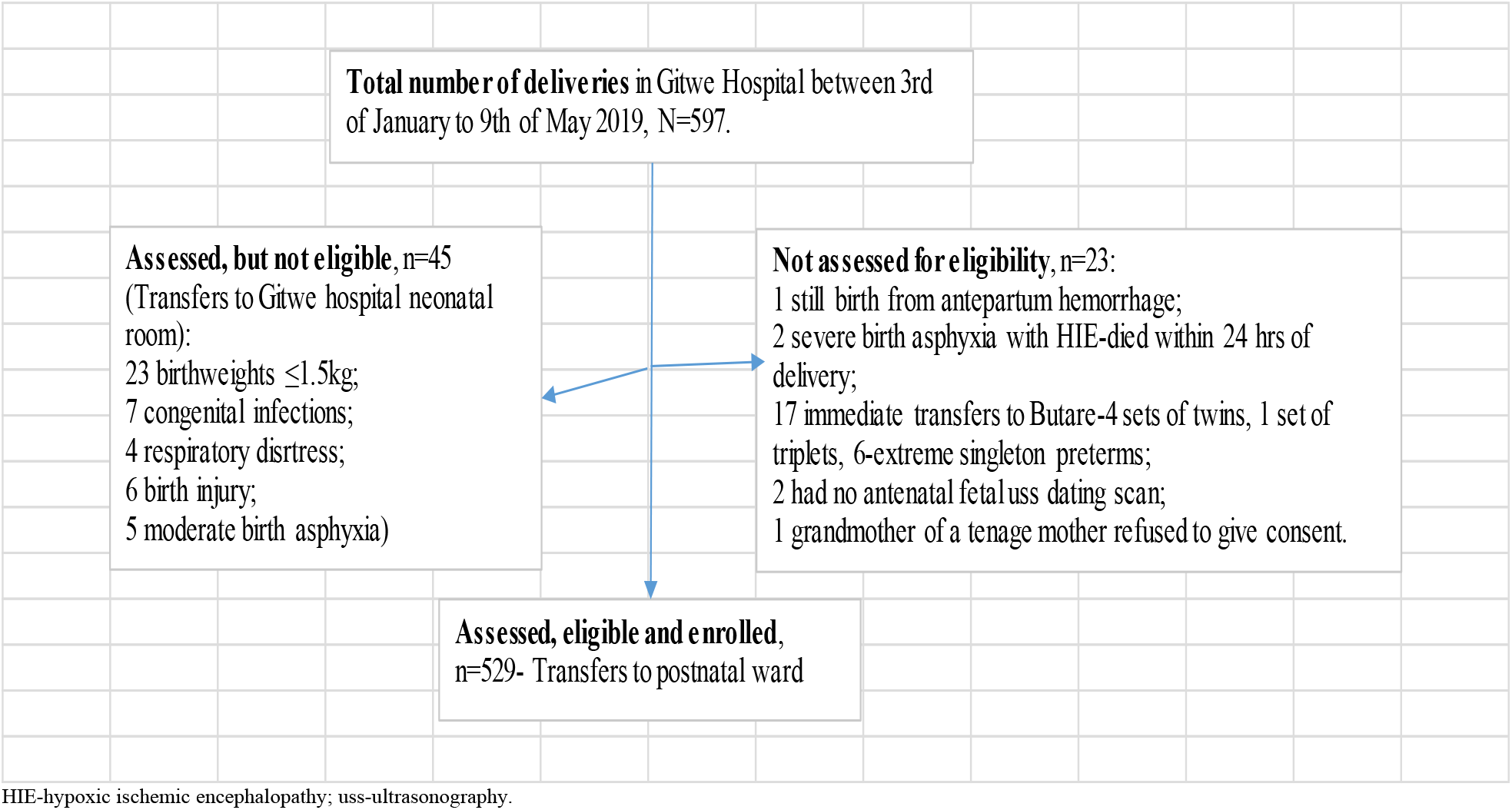
Flow of participants from admission to recruitment into study.

### Characteristics of postdate and term newborns in the study

Out of the 503 newborns analysed, postdate babies were 199 (39.6%), while the term babies were 304 (60.4%), Table 1. The range of gestational age for the postdate babies was 41-43 weeks, while for the term babies was 38-40weeks. The median gestational age and interquartile ranges for the postdate and term babies were 41 (IQR 41, 42 weeks) and 40 (IQR 39, 40), respectively. The mean birthweight of the postdate (3.20kg, SD 0.43) was slightly higher than that of the term babies (3.15kg, SD 0.49), but this difference was not significant, p=0.213.

**Table 1:** Characteristics of postdate and term newborns in the study, n=503.

|  |  | Gestational age at birth |  | Total |
| --- | --- | --- | --- | --- |
|  |  | Postdate (%) | Term (%) |  |
| **Birth size-GA** (p<0.001) | SGA | 69 (63.9) | 39 (36.1) | 108 (100.0) |
|  | AGA | 126 (34.5) | 239 (65.5) | 365 (100.0) |
|  | LGA | 4 (13.3) | 26 (86.7) | 30 (100.0) |
| **Sex** (p=0.631) | Female | 96 (40.7) | 140 (59.3) | 236 (100.0) |
|  | Male | 103 (38.6) | 164 (61.4) | 267 (100.0) |
| **Mode of delivery** (p=0.117) | Cesarean section | 68 (35.2) | 125 (64.8) | 193 (100.0) |
|  | Vaginal delivery | 131 (42.3) | 179 (57.7) | 310 (100.0) |
| **Birth weight** (p=0.116) | Low birth weight | 10 (24.4) | 31 (75.6) | 41 (100.0) |
|  | Normal birthweight | 184 (40.9) | 266 (59.1) | 450 (100.0) |
|  | High birthweight | 5 (41.7) | 7 (58.3) | 12 (100.0) |
| **Place of delivery** (p=0.762) | Outside health facility | 1 (50.0) | 1 (50.0) | 2 (100.0) |
|  | Within health facility | 198 (39.5) | 303 (60.5) | 501 (100.0) |
| **Total** |  | **199 (39.6)** | **304 (60.4)** | **503 (100.0)** |

Their intrauterine growth was the only characteristic that was significantly different between the term and postdate groups of babies. The overall prevalence of SGA birth was 21.4% (108 out of 503). 69 out of 108 (63.9%) SGA were in the postdate group, while 26 out of 30 (86.7%) LGA were in the term babies group, p<0.001. Both the postdate and term groups of babies were similar with regards to sex, mode of delivery, place of delivery and birthweight, p>0.05, Table 1.

### Characteristics of mothers of postdate and term newborns in the study

In Table 2, only maternal occupation (p=0.005), marital status (p=0.004) and MUAC (p=0.049) were significantly different in the 2 groups of newborns. A higher percentage of professional mothers (80.0%) gave birth after term, while a higher percentage of shop owners (81.8%) gave birth at term. A higher percentage of married mothers gave birth after term (41.3%), while 79.1 % of the unmarried mothers gave birth at term. 41% of mothers with high MUAC delivered after term, while 73.1% of thin mothers delivered at term.

**Table 2:**
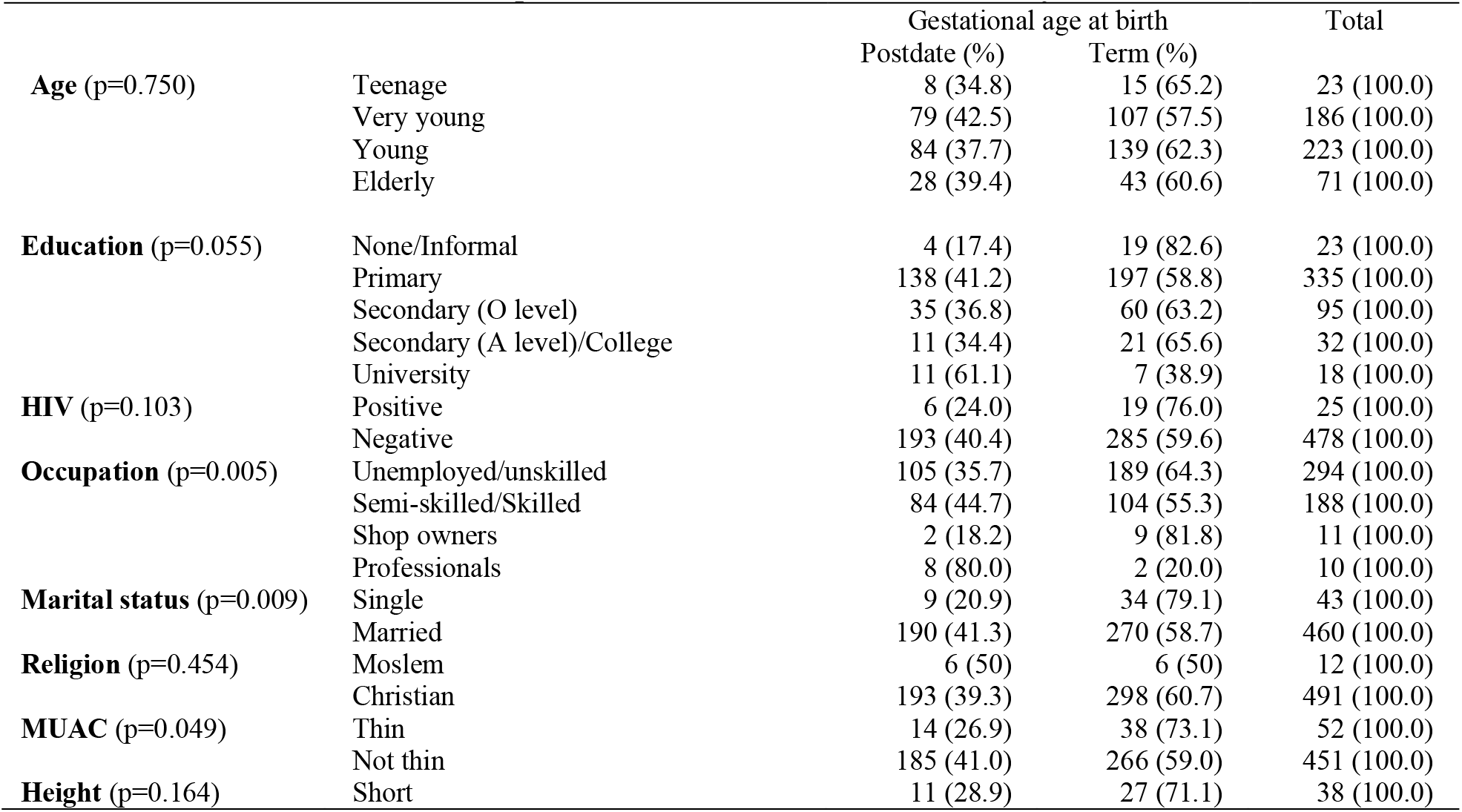
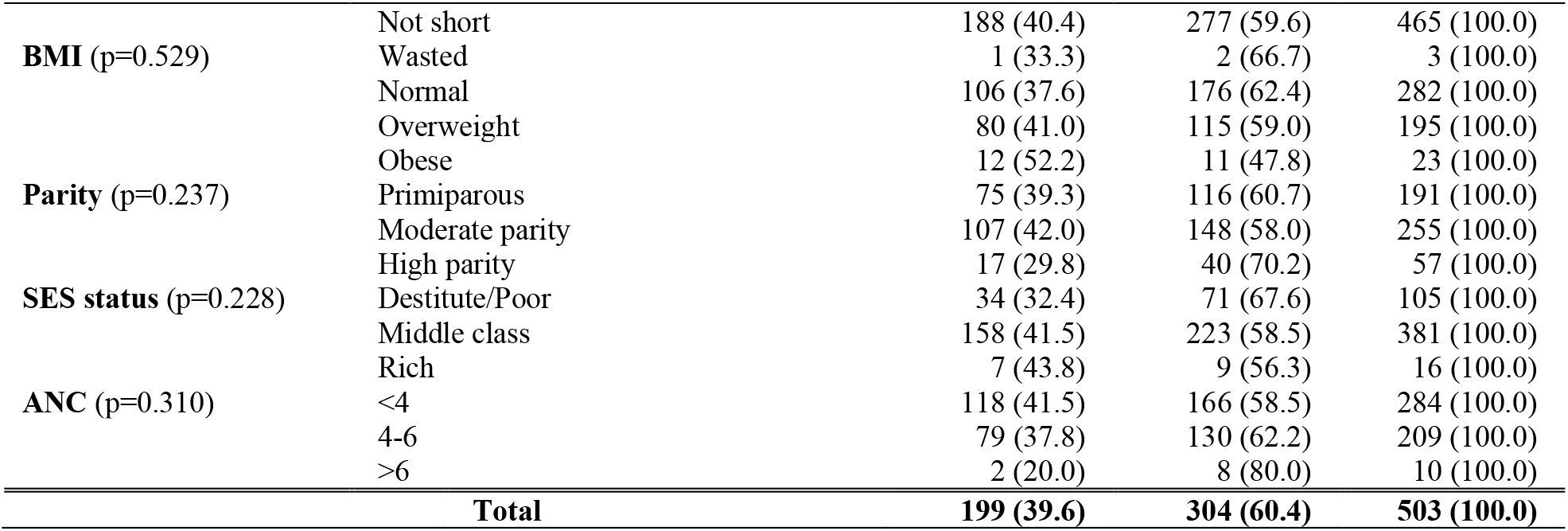
Characteristics of mothers of postdate and term newborns in the study, n=503.

### Environmental characteristics of postdate and term newborns in the study

In Table 3, use of water (p=0.001) and toilet facilities (p=0.004) located within or outside the home yard contributed significantly to the duration of gestation. A higher percentage of mothers, who used water (44.7%) and toilet (46.9%) facilities located outside the home yard delivered after term; while a higher percentage of mothers, who used water (71.4%) and toilet (65.8%) facilities located within the home yard delivered at term.

**Table 3:**
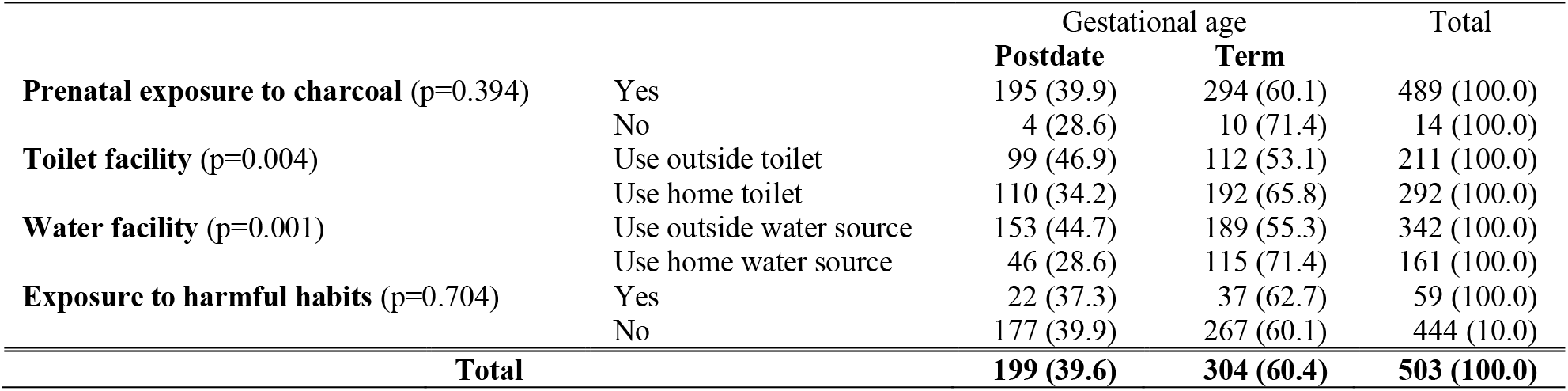
Environmental characteristics of postdate and term newborns in the study, n=503.

### Binary logistic regression analysis of factors predicting postdate pregnancies

Factors in Tables 1-3, with p-values <0.100 were included in binary logistic regression analysis-intrauterine growth size, maternal education, occupation, marital status, MUAC, use of toilet and water facilities. Only intrauterine growth status of the newborn at birth and maternal occupation remained significant predictors of postdate delivery. The AGA (p<0.001; OR=0.09; 95% CI 0.03, 0.30) and LGA (p=0.063; OR=0.35; 95%CI 0.12, 1.06) newborns were less likely to be delivered postdate compared to SGA babies. The confidence interval for maternal occupation was too wide (1.03, 156.14) to be precise, Table 4.

**Table 4:** Binary logistic regression analysis of factors predicting postdate delivery in the study, n=503.

|  |  | Wald | df | p-value | OR | 95% C.I. for OR |  |
| --- | --- | --- | --- | --- | --- | --- | --- |
|  |  |  |  |  |  | Lower | Upper |
| Step 1<sup>a</sup> | SGA | 34.588 | 2 | 0.000 |  |  |  |
|  | AGA | 16.001 | 1 | 0.000 | 0.09 | 0.03 | 0.30 |
|  | LGA | 3.461 | 1 | 0.063 | 0.35 | 0.12 | 1.06 |
|  | None/Informal | 3.857 | 4 | 0.426 |  |  |  |
|  | Primary (1) | 2.611 | 1 | 0.106 | 4.351 | 0.73 | 25.89 |
|  | Sec O (2) | 0.585 | 1 | 0.444 | 1.739 | 0.42 | 7.19 |
|  | Sec A/College (3) | 0.508 | 1 | 0.476 | 1.681 | 0.40 | 7.01 |
|  | Univ (4) | 1.320 | 1 | 0.251 | 2.443 | 0.53 | 11.21 |
|  | Unemp/Unskilled | 4.144 | 3 | 0.246 |  |  |  |
|  | Semi-skilled/Skilled (1) | 1.865 | 1 | 0.172 | 4.126 | 0.54 | 31.55 |
|  | Shop owners (2) | 1.591 | 1 | 0.207 | 3.577 | 0.49 | 25.93 |
|  | Professionals (3) | 3.931 | 1 | 0.047 | 12.680 | 1.03 | 156.14 |
|  | Married (1) | 3.850 | 1 | 0.050 | 2.273 | 1.00 | 5.16 |
|  | High MUAC (1) | 2.191 | 1 | 0.139 | 1.739 | 0.84 | 3.62 |
|  | Toilet within (1) | 0.909 | 1 | 0.340 | 0.792 | 0.49 | 1.28 |
|  | Water within (1) | 0.204 | 1 | 0.652 | 0.875 | 0.49 | 1.56 |
|  | Constant | 0.019 | 1 | 0.891 | 0.872 |  |  |

In a multinomial logistic regression analysis of postdate and term deliveries predicting fetal intrauterine growth retardation, the odds of a postdate pregnancy producing a SGA newborn was 11.05 (95% CI 3.74, 35.57) compared to a term delivery.

### Distribution of SGA babies among the gestational ages in the study

In Table 5, the percentage of SGA babies increased from 14.3% at 38 weeks to 80.0% at 43 weeks gestation, p<0.001. On the contrary, the percentages of LGA was dropping as the gestation prolongs. Of note, at 42 and 43 weeks gestation, there was no LGA baby. The percentage of AGA was highest at 40 weeks gestation (85.6%) and thereafter declined. The differences in SGA distribution across the gestational age was significant (p<0.001).

**Table 5:**
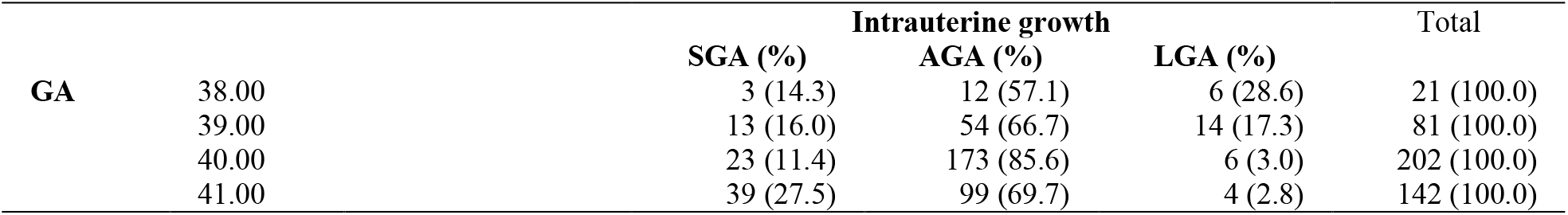
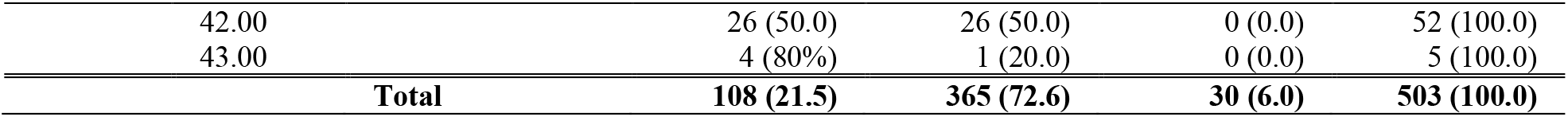
Distribution of SGA babies across the gestational ages in the study, n=503, p<0.001.

In Table 6, 66 out of the 69 (95.7%) postdate SGA babies were symmetrical SGA, while 30 out of the 39 (76.9%) term babies were symmetrical SGA, p=0.003; OR=6.60; 95% CI (1.67, 26.1). The postdate babies were 6.6 times more likely to be symmetrically grown SGA than their term counterparts.

**Table 6:** Distribution of SGA types accross the gestational ages in the study, n=108, p=0.003; OR 6.60.

| Gestation |  | Type of IUGR (%) |  | Total |
| --- | --- | --- | --- | --- |
|  |  | Symetrical<br>(PI>2) | Asymetrical<br>(PI≤2) |  |
|  | Postdate | 66 (95.7) | 3 (4.3) | 69 (100.0) |
|  | Term | 30 (76.9) | 9 (23.1) | 39 (100.0) |
|  | **Total** | **96(88.9)** | **12(11.1)** | **108(100.0)** |

### Breastfeeding initiation and prelacteal feeding amongst the postdate and term babies in the study

Among babies, whose first breastfeed was delayed beyond 1 hour, 17 out of 79 (21.5%) were postdate, while 62 out of 79 (78.5%) were term babies (p<0.001; OR=0.37; 95% CI 0.21, 0.65). The postdate babies had 63% lower risks of experiencing delayed first breastfeed than their term counterparts. Among babies, who were offered prelacteal feeds, 12 out of the 32 (37.5%) babies were postdate, while 20 out of the 32 (62.5%) babies, were term babies, p=0.805.

## Discussion

It is assumed that the postdate baby is a rare occurrence in today’s neonatal practice, because mothers have become more pro-active in expressing concerns about delivering past their due dates.^3, 19^ However, this assumption is defeated by the high rate (39.6%) of postdate deliveries (GA>40 weeks) observed in this high-altitude study.

The postdate babies were more likely to have experienced intrauterine growth retardation compared to their term counterparts, and the risk progressively increased with advanced gestation, from 14.3% at 38 weeks to as high as 80% at 43 weeks gestation. The rate of AGA babies peaked at 40 weeks gestation and then declined afterwards. The postdate babies were also 6.6 times more likely to have developed type 1 fetal growth retardation (symmetrical SGA) than their term counterparts. Symmetrical SGA suggest a decompensated state, from chronic, early onset or severe intrauterine insult to the fetus. The symmetrically growth retarded newborn has more serious short and long-term complications than the asymmetrically grown SGA newborn. Given these results, it would be prudent to induce labor at 40 weeks rather than later, in low-income, high-altitude settings, such as in this study.^6^

It is anticipated that native highland people, who have been historically subjected to hypobaric hypoxia for decades, undergo selection for beneficial genetic variations,^20^ that might contribute to partial mitigation of high altitude effects on fetal growth, as has been documented in such populations.^21^ This mitigation effect is not apparent in this current study. This might suggest that prolonged residence in high altitude location combined with other adverse maternal and environmental factors, could be responsible for this non-compensated adverse neonatal outcome picture. The morbidities and mortalities associated with symmetrical SGA are higher than those experienced with the asymmetrical SGA. The SGA rate in this current study was 21.4%, 4 times higher than the preterm birth rate. This suggest that the emphasis and focus of intervention should be on SGA in high altitude areas.

Comparing this current study findings with those of Dolma et al. (2022),^22^ who prospectively studied 316 high-altitude pregnant women (in Ladakh, 3524 m-twice higher than current study site); prevalence of SGA was 14% in Dolma et al’s study, lower than the 21.4% of this study, which is contrary to expectation. Of note the mean birthweight of newborns in both studies were similar, emphasizing the need to classify all newborn, especially in this high risk areas, in order to identify and manage the SGA problems effectively.^23^ Maternal characteristics in both studies may have contributed to these unexpected differences. Both studies had equal percentages of primiparous mothers (38%), however, the percentage of multiparous mothers in the Dolma’s study was at least 5 times higher than this current study (62% vs 11.3%). Also, the mothers in the Dolma study were older (mean age 28.4years vs 27.8years), more educated, with better employed and higher socioeconomic status than in this current study. The poor environmental conditions (water and toilet facilities) in this current study may have also contributed to the higher prevalence of SGA.

Grant et al (2021)’s global systematic review with meta-analysis of 59 studies and 1 604 770 pregnancies; observed that there was an increased risk of LBW, SGA, spontaneous preterm births by 47%, 88% and 23% respectively, in populations at high altitudes compared to those at low altitudes.^9^ Levine et al’s retrospective cohort study of 550,166 women in low (0-1999m), moderate (2000-2900m) and high altitude (3000-4340m) residences reported a preterm birth rate of 5.9% (comparable to the 4.9% preterm of current study) with no significant difference in preterm birth rate across the 3 altitudes.^24^ This evidence also point to SGA (rather than preterm birth) as a priority focus for intervention in high altitude areas.

Unlike in Kandalgaonkar et al’s study,^5^ there was no significant difference in cesarean section delivery between postdate and term deliveries in this current study. High MUAC and not BMI was a predictor of postdate pregnancy in this current study, unlike in Stotland’s study.^13^ Moderate parity and female gender (as in earlier literature), rather than primiparity and male gender predisposed to postdate pregnancy in this study.^3^Of note, mothers that infrequent antenatal clinic were predisposed to postdate pregnancy in this study. This is similar to Deng et al’s report that stated that the risk of postterm births decreased as the number of antenatal visits increased in retrospective study of 6,240,830 mothers in China.^25^

One of the limitations of the study, was the challenge of accurately determining the gestational age of the babies, as mothers may have difficulties in recalling their LMP and registering late for their fetal ultrasound dating. These challenges were mitigated to some extent by ensuring all the mothers had at least a dating scan before delivery, as is the policy in Rwanda and combining it with mother’s report on her LMP and/or investigator’s ballard examination estimate. Another limitation was the exclusion of very small and sick babies from the study, (because of prioritising resuscitation and transfer for specialist care in another health facility), which may have reduced the power of the study.

The strength of the study include, the comprehensive assessment and classification of the newborns that took place during the study, which was an uncommon practice in the non-specialised health facilities.

## Conclusion and Recommendations

The rates of postdate and SGA births were unexpectedly high for the level of altitude. Unlike in low land studies, where prolonged pregnancies were associated with macrosomia, postdate deliveries in this high-altitude population, were more commonly associated with SGA birth and their complications. The strong association of symmetric SGA (rather than asymmetric SGA) with postdate pregnancies suggests longstanding intrauterine insult from probably combined effects of chronic hypoxia from high altitude residence and maternal factors. The authors of this study strongly recommend that high altitude pregnancies should not be allowed to progress beyond 40weeks gestation and more resources should be committed to identifying high risk pregnancies with tendencies for prolonged pregnancies, improve maternal nutrition and oxygen status during pregnancy. Maternal and public education on the benefits of adequate antenatal visits and care to facilitate early detection of postdate pregnancy and intervention. Though, their birthweights were preserved to some extent, even at high altitude, the newborns should be classified at birth, their problems anticipated and managed appropriately for an improved neonatal care, survival and wellbeing.

## Data Availability

All data produced in the present study are available upon reasonable request to the authors

## Author Contributions

The corresponding author (Dr Adenike Oluwakemi Ogah) conceived and designed the study, collected data and conducted data analysis, interpreted the results, and drafted the manuscript. James Aaron Ogbole reviewed and edited the manuscript for intellectual content. All the authors approved the final manuscript for submission.

## Acknowledgements

The authors are extremely grateful to the participants involved in this study, to the staff of Gitwe Hospital and clinic in Rwanda and to the research team. I am also indebted to my supervisors (Prof Andre Venter and Prof Corina Walsh), and my statistician Prof Gina Joubert, who analysed all the data for my PhD thesis.

